# The European Reality Regarding ICU Follow–Up Clinics: A Systematic Review

**DOI:** 10.1101/2023.06.24.23291854

**Authors:** Marios Charalampopoulos, Panagiota Triantafyllaki, Christina-Athanasia Sampani, Christos Triantafyllou, Dimitrios Papageorgiou, ICU Follow-Up Care Lab

## Abstract

**Introduction:** The ICU Follow up Clinics support and treat adult patients in the community in order to cover the needs that occur after their discharge from the ICU at the hospital.

**Aim:** To study the effectiveness and the way of the ICU Follow up Clinic operation management for adult patients after their discharge from the hospital.

**Methods:** A search was performed on the Greek and international literature, as well as at the online Databases PubMed, Cochrane, EMBASE and Google Scholar. Exclusion and integration criteria were set for the studies found, and a flow chart was created for the studies included.

**Results:** Through the search, 30 articles were found matching the subject under study, and after further evaluation, seven articles were included. The majority of the articles highlight the importance of these services at the patients’ follow-up after discharge. More specifically, through these services, the mortality rate, the risk of recurrence, and the readmission risk are decreased as well as the patient’s mental health is improved.

**Conclusions:** The main aim of these Services is the follow-up of the adult patients after discharge, the consultation, and the application of therapeutic protocols in order to face the primary pathological cause, as well as matters that occurred as a result of the hospitalization. At the same time, they can help to detect the needs of the patients that occurred after the discharge so that finally, improve the care given and the patient’s outcome.

## Introduction

The Intensive Care Unit (ICU) is defined as the hospital’s Department where patients, who need intensive and immediate support of their vital functions to be kept alive, are treated. The purpose of the ICU is the best possible care that will help the patient survive and return to the pre-illness state with the least possible complications and disabilities. The ICU Patient Monitoring Services were created in order to orientate the care of the patients towards a holistic character. These services are mainly composed of nurses and doctors that support the needs that arise in new daily life in the community after an ICU stay with the patient.^1,2,3,4,5^

The first published reference to these Services was observed in 1997 in London, England, while already in 1985, there were references to corresponding health facilities in the United Kingdom. Later in 1989, another was created another Service by the King’s Fund Panel and in 1999 by the Audit Commission.^6^

Structurally, the Follow-up Services are mainly composed of doctors and nurses, while the team also includes other health professionals such as physical therapists, psychologists, etc. Operationally, there is no specific mode of operation, but it is adapted to the circumstances according to the needs of the population it is addressed. The main purpose of these Services is to monitor the patient’s progress after hospital admission, counseling, and the application of therapeutic protocols to cope with the consequences arising after ICU hospitalization (impaired mobility, mental health, physical endurance).^1-6^

## Methods

We collected data from the international electronic scientific bases PubMed, SCOPUS, and EMBASE as well as at the Greek IATROTEK-online, both in English and Greek language, respectively. No limitations were applied regarding the articles’ publication time. International or local locations of survey appliances were included. As keywords were used, the following: “critically ill patient”, “after ICU complications”, and “ICU follow-up services”.

## Results

In the absence of Greek literature, this article is based mainly on English studies without time limitations. The articles that met the inclusion criteria and were inserted into this systematic review were seven. The above results and the flow chart of the studies included are analyzed below (Figure 1).

**Figure 1:**
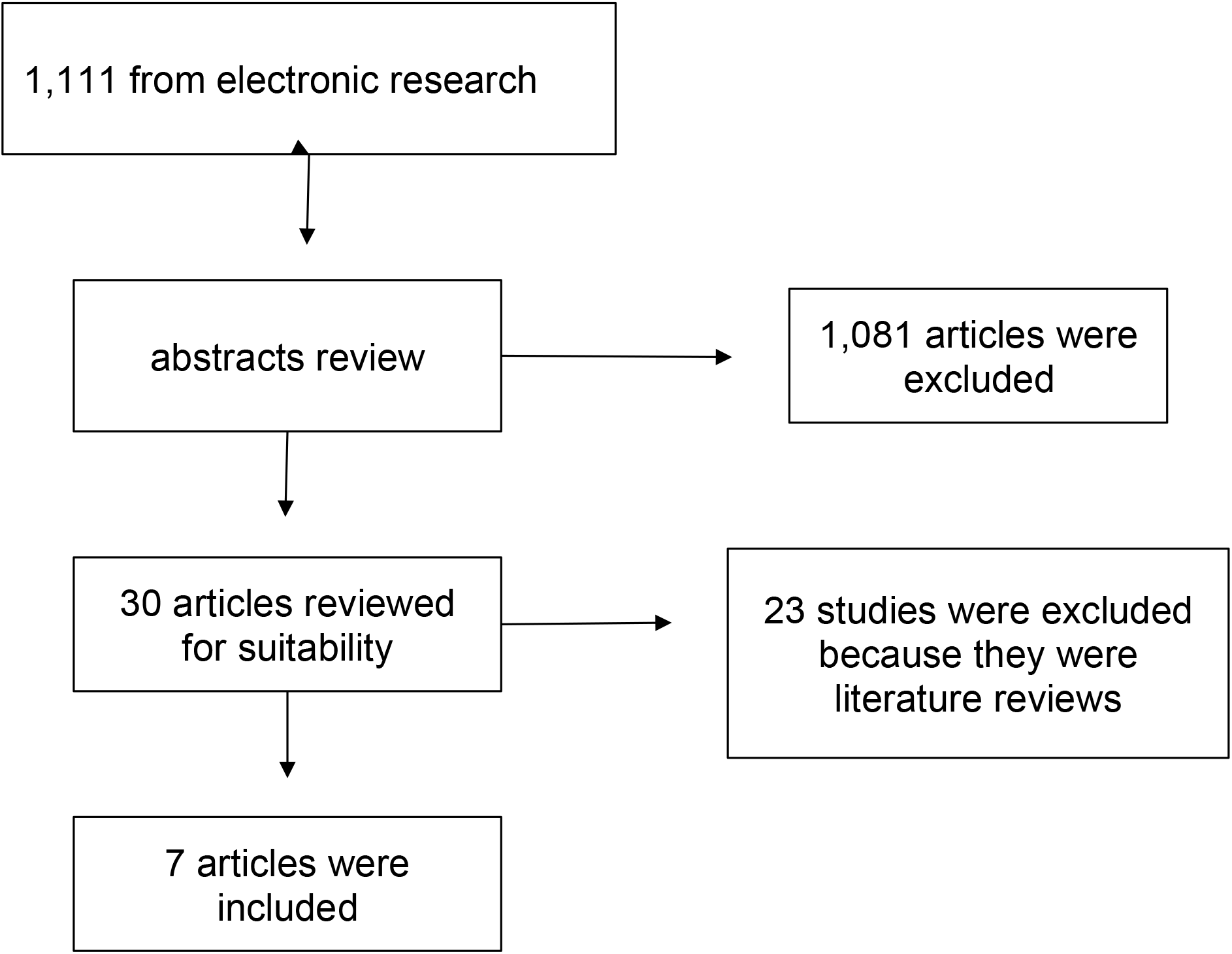
Flow chart of the systematic review.

**Table 1.**
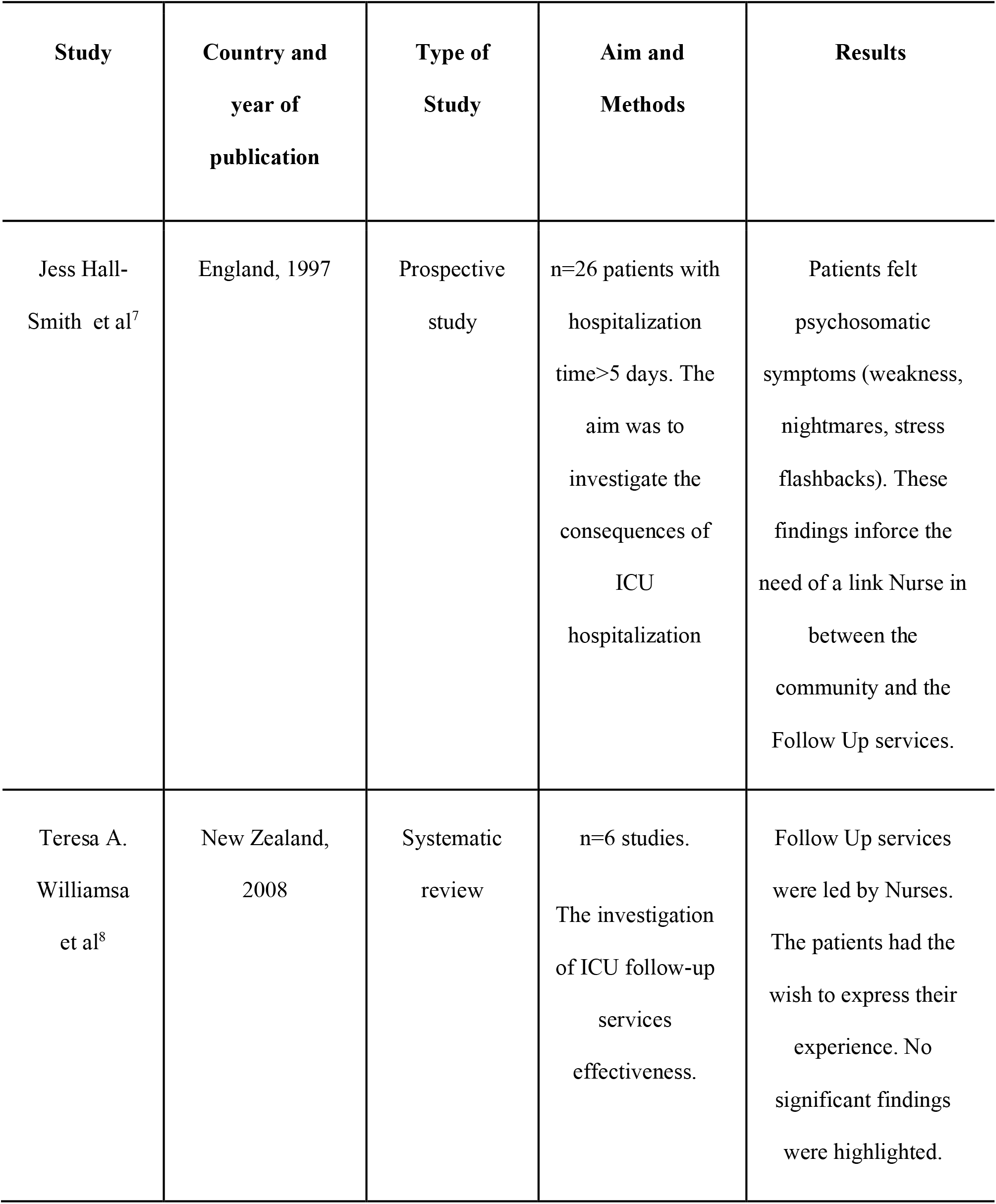
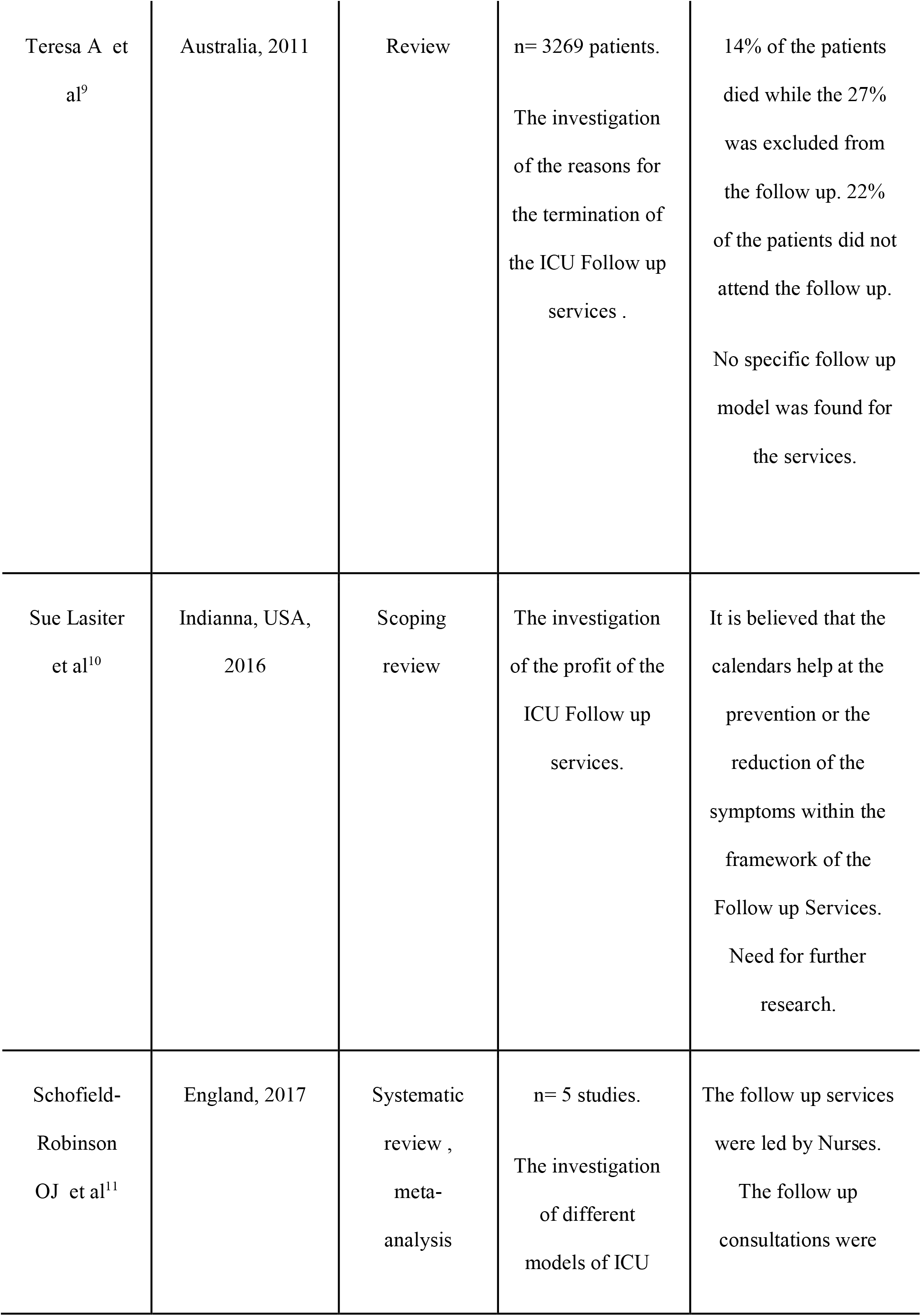
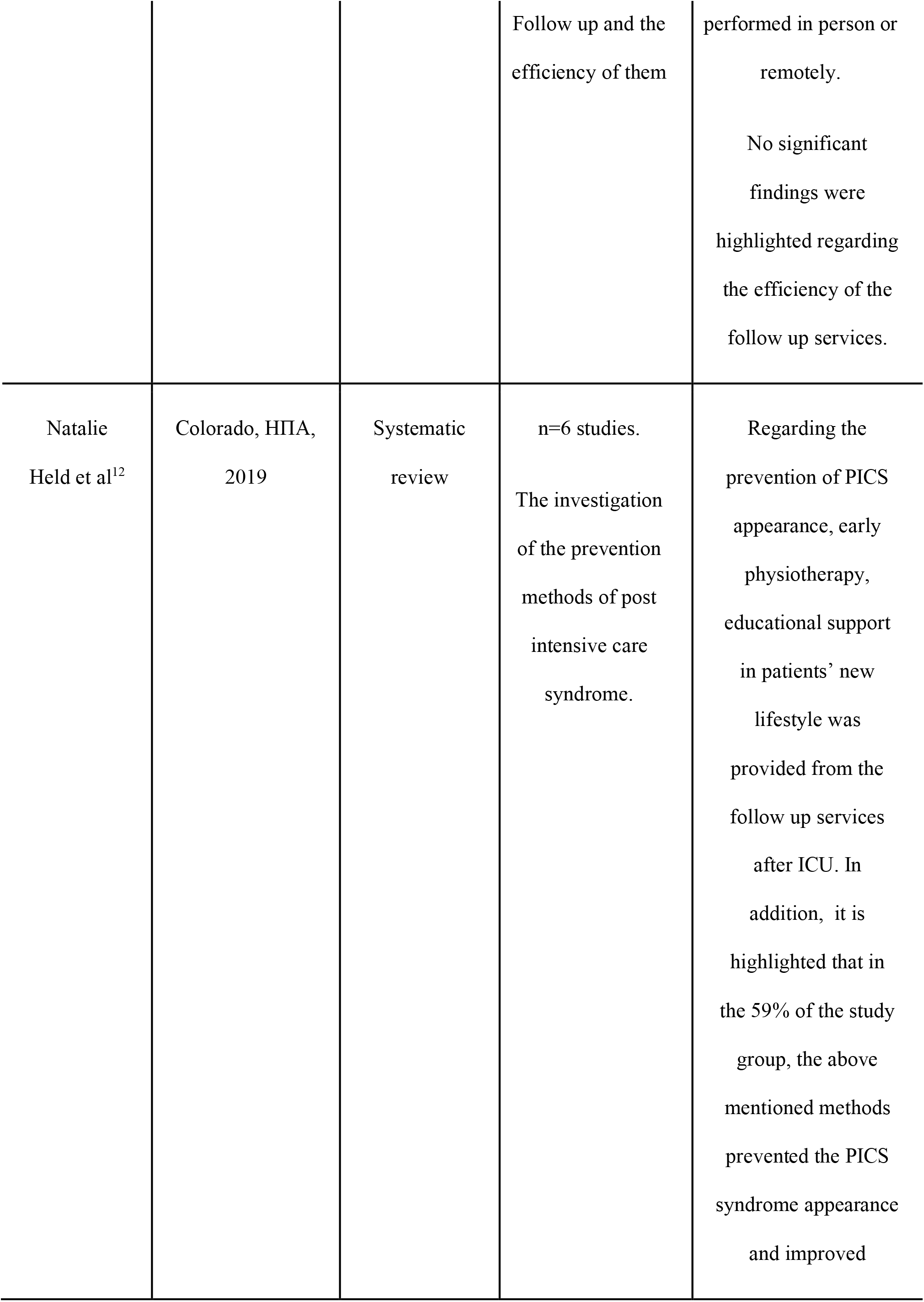
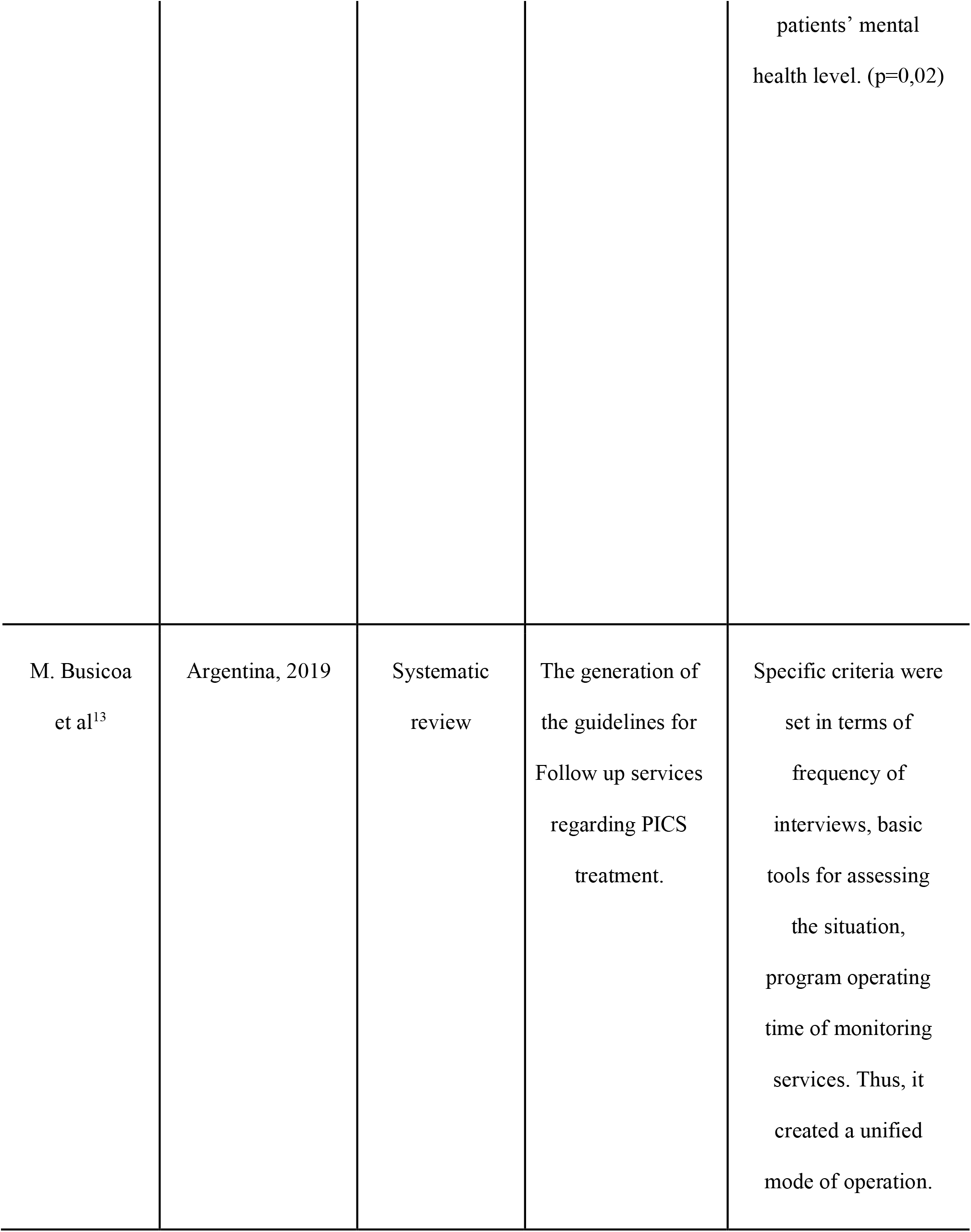
Summary of the studies in the systematic review

### Jess Hall-Smith et al. (1997)^7^

The prospective study aimed to investigate the consequences that occur after ICU hospitalization according to ICU Follow-Up services findings. They included 26 studies in which the patients were hospitalized in the ICU for more than five (5) days. In addition, further limitations were applied regarding the patients’ comorbidities in order to facilitate the follow-up procedure. Following these limitations, HIV, final-stage cancer, degenerative diseases, cardiovascular disease patients, and patients followed at the same time by other medical teams were excluded. At the results, patients were found experiencing psychosomatic symptoms (weakness, nightmares, stress, flashbacks), which enforces the need the link Nurses between community and ICU Follow-Up services.

### Teresa A. Williams et al. (2008)^8^

This systematic review aimed to investigate the impact of ICU Follow-Up services on ICU patients after hospital discharge. Electronic research was performed in international scientific databases where six articles were included. Regarding the first follow-up appointment since the hospital discharge, it was defined as between two (2) to three (3) months. The Follow-Up services were led by Nurses. As for the results, the patients had the wish to express their experience, but no significant findings were highlighted.

### Teresa A et al. (2011)^9^

This review aimed to investigate the reasons for the termination of the ICU Follow-Up services. From the cyber research that was performed for the 2006-2010 period of time, ten (10) studies and 3269 patients in total were included. Of the total number of patients, 14% died, while 27% were excluded from the follow-up. Regarding the continuity of follow-up, 22% of the patients did not complete the program. Furthermore, it is highlighted that the follow-up services have a positive impact on the prevention of the complications that occur in the community as a result of the previous ICU hospitalization and, in addition, on hospital readmissions.

### Sue Lasiter et al (2016)^10^

This scoping review aimed to investigate the profit of ICU Follow-Up clinics. It is believed that the calendars help prevent or reduce symptoms within the framework of the Follow-Up Services. There is a great need for further research, though.

### Schofield-Robinson OJ et al. (2017)^11^

The main goal of this meta-analysis was the investigation of the efficiency of Follow-Up ICU services and the different possible models of operation. Five (5) studies were included. The Follow-Up services were led by Nurses. The Follow-Up consultations were performed in person or remotely within a week, a month, or six months. For the consultations, the HRQoL questionnaire was used. No significant findings were highlighted regarding the efficiency of the Follow-Up services.

### Natalie Held et al (2019)^12^

The aim of this systematic review was the investigation of the prevention methods for post-intensive care syndrome. Six (6) studies were included. Regarding the findings, various methods were performed, for example, early physiotherapy, mobility rehabilitation and new patients’ lifestyle educational support reflecting after ICU hospitalization consequences. In addition, it is highlighted that in 59% of the study group, the above-mentioned methods prevented the PICS syndrome appearance and improved patients’ mental health status (p=0.02).

### M. Busicoa et al (2019)^13^

The aim of this systematic review was the generation of guidelines for Follow up services regarding PICS treatment. Regarding the first appointment after discharge, there was an option within one (1) month in order to detect any consequences early or within 12 months for the less critically ill patients. For this purpose, the APACHE II score was used, with a desired score of more than 14 combined with the duration of the mechanical ventilation, less than seven (7) days. Furthermore, the duration of the ICU hospitalization, in order to admit at the Follow-Up services, was set at more than ten (10) days. Also, the presence of a psychiatric disorder or impaired knowledge level was considered an exclusion criterion. In conclusion, the frequency of the interview and the available consultation tools defined the guidelines.

## Discussion

Based on the surveys, a variety of results and conclusions were observed. For example, in terms of the minimum length of stay in the ICU in order to be included in the services, some surveys set a limit of only 24 hours, such as the European Society for Intensive Care, while others, such as the Homerton Hospital survey, set a limit of five (5) days due to the high number of annual ICU admissions.

Regarding the criteria for inclusion in the follow-up services, the Swedish study set the criterion of language (proficiency in Swedish), while other studies, such as Daffurn et al. and Homerton Hospital, set the criterion of distance from the clinic to include only patients who were in close proximity. In contrast to the above, in the Argentinean study, qualitative criteria such as the APACHE II prognosis scale with a desired score above 14 and a number of other clinical ‘parameters’ were set. Regarding the application of follow-up, the “patient diary” method was observed in the Swedish study in order to have a complete record of events from the ICU hospitalization and to observe any progress in the future, while in the majority of studies, the assessment of the patient’s psychological and physical condition was performed using the SF-36, EQ-5D questionnaires.

Regarding the time of follow-up of patients, there was a wide variation as some studies reported a time of only six months, while others reported up to 5 years. This is justified by dynamic parameters such as patient survival time and place of residence, which may significantly limit the follow-up time. In terms of the evaluation of the above, the diary-based follow-up model, both in the Swedish study and in the study by Combe et al. and Haraldsson L, is satisfactory to patients, but this is not proven by qualitative studies.

Regarding other methods, such as the application of quality-of-life assessment questionnaires alongside therapeutic interventions by the medical team, the patient benefit was observed in the areas of mental and physical health. This fact is confirmed in the studies of Lasiter S (2017), Modrykamiem A et Al (2012), Karaldsson L (2015), and others. Regarding costs, no research has shown a benefit to the hospital. However, in a study by Teresa A et al., it was reported that follow-up services prevent complications arising in the community that would likely cause readmission to the hospital and, thus, the additional financial burden on the Health System. As for the staffing of the Monitoring Services, the literature search revealed a diverse group of health professionals. However, Monitoring Services may be managed by a nurse or a physician, or a combination of both.

Finally, for a variety of reasons, these structures face obstacles that they have to overcome in order to continue operating, one of the main ones being of funding. However, regarding

Monitoring Services, no research data demonstrates an economic benefit to the hospital that creates the Service. Consequently, apart from the health interest, there are no supporting arguments for funding such programs for the purpose of generating revenue. At the same time, even from a health perspective, there are insufficient research results to demonstrate the substantial benefit of the program to patients. Also, comparing the needs in the community of patients who were hospitalized only in Nursing Departments with patients who were hospitalized in ICUs, there is no significant difference, although as patient categories, they are completely different in terms of severity. In addition to the above, an obstacle to the functioning of monitoring services may be the ‘object’ of monitoring itself, the patients. It was mentioned earlier that the main model for monitoring patients is face-to-face communication through meetings. However, this is not always possible as operational issues such as distance arise. A hospital may be treating people from wider areas of greater distance, and, as a result, these patients may not be able to travel as often as required for follow-up appointments.

In addition, in many cases, a relapse of the patient’s condition causes him to abstain from meetings even if he is benefiting from them. In a different category of patients, there is a refusal to participate in follow-up programs, possibly due to fear of the hospital environment and the associated memories, but also due to a false belief that they do not need follow-up as they have finally overcome their health issues.

### Conclusions

For the best possible provision of care to patients, it is beneficial to try to create guidelines on the operation of the Monitoring Services so that there is uniformity in the way organization and operation always adapt to the needs of the population and the needs of τhe target population. Regarding the Greek reality, the absence of such a comprehensive structure contributes to the increase in hospitalization costs, hospital-related infections, and, finally, the absence of holistic care for this category of patients. In conclusion, the follow-up services function as a tool to support patients in the community where the risk of relapse and readmission is characterized as quite likely. In addition, it is possible to be used as a research tool to identify the needs that arise after an ICU hospitalization in order to further improve the care provided and, ultimately, to improve the quality-of-care outcome of patients (therapeutic protocols, technological equipment, better rehabilitation).

## Data Availability

All data produced in the present study are available upon reasonable request to the authors

## Notes

### Competing Interest Statement

The authors have declared no competing interest.

### Funding Statement

This study did not receive any funding

## Bibliography

1. Valentin A, Ferdinande P; ESICM Working Group on Quality Improvement.Recommendations on basic requirements for intensive care units: structural and organizational aspects. Intensive Care Med. 2011, 37(10),1575–87.

2. Wunsch H, Gershengorn H, Scales DC. Economics of ICU organization and management. Crit Care Clin. 2012,28(1),25–37.

3. Central Health Council of Greece - Minimum requirements for the operation of Intensive Care Units (n.d). Available from: https://www.icu.gr/DOCS/2017/2017_6_2016.pdf (6/2020)

4. Priorities for admission and discharge criteria for patients in adult intensive care units (n.d). Available at: http://www.icu.gr/DOCS/2017/2017_05_29.pdf (6/2020)

5. Calliope P., Organization and Management of Intensive Care Unit, online, 2017, 59–75. Available at: http://dione.lib.unipi.gr/xmlui/bitstream/handle/unipi/10492/Michou_Kalliopi.pdf?sequence=1&isAllowed=y (6/2020)

6. Karanikolas. Ethical Dilemmas at the I.C.U. n.d, 1–6 Available at: https://eclass.upatras.gr/modules/document/file.php/MED1029/BioethicsKaranikolas.pdf (6/2020)

7. Hall-Smith J, Ball C, Coakley G, Follow-up services and the development of a clinical nurse specialist in intensive care, ICC Nursing, 1997, 13, (5), 1997, 243–248.

8. Teresa A. Williams A, Gavin D. Leslie B. Beyond the walls: A review of ICU clinics and their impact on patient outcomes after leaving hospital. Aust Crit Care 2008, 21(1), 6–17.

9. Teresa A. Williams, Gavin D, Leslie. Challenges and possible solutions for long term follow-up of patients surviving critically. Aust Crit Care 2011, 24(3), 175–85.

10. Needham d, Davidson J, Cohen H et al. Improving long-term outcomes after discharge from intensive care unit: Report from a stakeholders’ conference. Crit Care Med 2012, 40(2), 502–9

11. Schofield-Robinson OJ, Lewis SR, Smith AF, McPeake J, Alderson P. Follow-up services for improving long-term outcomes in intensive care unit (ICU) survivors (Protocol). Cochrane Database Syst Re 2018, 11(11), CD012701.

12. Held N, Moss M. Optimizing Post-Intensive Care Unit Rehabilitation. Turk Thorac J 2019, 20(2), 147–52.

13. Combe D. The use of patient diaries in an intensive care unit. Nurs Crit Care Jan-Feb 2005, 10(1), 31–4

